# Unsupervised Tissue Concepts for Explainable Sarcoma Subtype Prediction from H&E

**DOI:** 10.64898/2026.05.15.26353333

**Authors:** Tom Bisson, Davis Ingram, Samyukta Singh, Annie Li, Samantha Flynn, Wei-Lien Wang, Albert E. Kim, Christopher P. Bridge, Elizabeth G. Demicco, Antonio Sorrentino, Sizun Jiang, Yin P. Hung, Alexander J. Lazar, A. John Iafrate

**Author notes:** **Authors’ disclosures** AJI is a founder, consultant and holds privately held equity in Monimoi Therapeutics and receives royalties from ArcherDx/IDT.

## Abstract

Soft tissue sarcomas are a rare, heterogeneous group of tumors whose diagnosis remains challenging because of overlapping morphology and limited access to sarcoma-specialized pathologists. Although pathology foundation models have shown promise in computational pathology, their clinical translation remains limited by insufficient interpretability, particularly in diagnostically complex settings such as sarcoma diagnosis. Here, we developed and evaluated an H&E-based AI framework for sarcoma subtype classification that focused on explanability. Using the CONCH v1.5 foundation model, we computed embeddings from a tissue microarray cohort of 2,545 cases spanning 19 sarcoma subtypes and trained an attention-based multiple-instance learning model that achieved a balanced accuracy of 77.38% ± 1.88.

To move explainability beyond attention-based localization, we trained a sparse autoencoder on patch-level embeddings to learn 768 recurring visual concepts. 90 high-activation concepts were reviewed by three senior pathologists and curated into morphologically meaningful and non-meaningful categories, yielding a semantic dictionary of 41 diagnostically relevant tissue concepts. We then trained a linear attention-based model on the 768-concept vectors, which retained much of the performance of the raw embedding-based ABMIL model, achieving a balanced accuracy of 73.74% ± 1.30. When restricting the linear model to pathologist-curated morphologic concepts only, balanced accuracy further decreased to 67.04% ± 1.27, suggesting that the residual performance gain in the full concept model was driven by inconsistent, technical, or diagnostically irrelevant concepts. Concept-level explanations of the curated linear attention-based model aligned with known sarcoma morphology, including lipogenic, myxoid, spindle-cell, pleomorphic, vascular, small round blue cell, and matrix-forming patterns, and reproduced patterns of diagnostic overlap observed in human sarcoma pathology.

Together, these results show that H&E-based foundation-model representations capture meaningful diagnostic structure within the known limitations of H&E in sarcoma diagnostics, but that their clinical value depends on whether this structure can be made interpretable to pathologists. Sparse autoencoder-derived concepts can address this critical gap by converting embedding-level signal into recurring morphologic patterns that pathologists can review and name, providing the foundation to link these patterns to subtype predictions. In doing so, this approach turns concept discovery into a practical form of diagnostic explanation, while also revealing where model performance is supported by recognizable histopathology and where it relies on diagnostically irrelevant or inconsistent visual patterns.

## Introduction

Soft tissue sarcomas (STS), a rare, heterogeneous group of over 100 histological subtypes with diverse clinical presentations [1], represent ∼1% of adult cancers, but up to 20% of pediatric cancers. Diagnostic precision in STS, although essential [2], remains challenging, with 20–25% disagreement among expert pathologists, often leading to inappropriate or delayed therapies and poorer outcomes [3]. These challenges are exacerbated by the limited availability of sarcoma-specialized pathologists [4], small biopsy sample sizes, and the expense of molecular diagnostics [5]. Diagnostic variability in STS remains a major source of concern: a 2022 multi-institutional review reported full agreement in fewer than half the cases examined, with major discrepancies in over 20% of them [6]. At present, STS diagnosis relies on hematoxylin and eosin (H&E) staining, conventional immunohistochemistry (IHC), molecular diagnostics, and expert interpretation [7]. This complex and extremely challenging workflow is time-consuming, tissue-intensive, and subject to significant inter-observer variability, rendering diagnostic consistency and scalability ongoing unmet needs, particularly in community hospitals and struggling health systems [8].

Sarcoma subtypes are morphologically well described, yet they rely on a limited set of architectural and cytological features that recur across diagnostic categories. Many of these features similarly recur across subtypes, including spindle cell fascicular growth, epithelioid or polygonal cell morphology, small round blue cell cytology, pleomorphic and multinucleated tumor cells, myxoid stromal change, lipogenic differentiation, vasoformative architecture, necrosis, and variable mitotic activity [1]. Some features remain relatively specific, such as plexiform capillary networks in myxoid liposarcoma, or osteoid deposition in osteogenic sarcomas, whereas others are shared across multiple diagnoses [9, 10, 11, 12]. Liposarcomas, for example, range from well-differentiated tumors with mature adipocytes and atypical stromal cells to myxoid variants with capillary networks and round cell progression [13, 14]. Gastrointestinal stromal tumors (GIST) span spindle-cell, epithelioid and mixed morphologies, sometimes with eosinophilic collagenous extracellular material [15], while PDGFRA-mutant GIST cases are enriched for epithelioid cytology and myxoid stroma [16, 17]. More generally, sarcoma diagnoses rely on distinct combinations of recurring morphologic features. Despite these characteristic features, a definitive classification often requires the integration of morphological, immunohistochemical and molecular findings.

Computational pathology has advanced rapidly [18], but the challenges in explaining model decisions in pathology have to date hindered large-scale clinical adoption [19]. AI solutions have successfully shifted from proof-of-concept tumor classifiers to assistive products and clinical-trial tools, but they still focus on high-volume, visually well-defined tasks: lesion detection in prostate biopsies, metastasis detection in lymph nodes, or standardized trial endpoint scoring. While the recent rise of pathology foundation models further expands transfer learning and multimodal reuse [20, 21, 22, 23], that progress has yet to translate into broadly accepted AI-assisted diagnostic workflows in routine pathology practice [18]. This lag in part stems from explainability methods inadequate to the demands of pathology. Specifically, most explainability methods currently rely on saliency or attention maps – useful for localizing case-level relevant image regions and providing prediction probabilities, but incapable of conveying semantic meaning or cross-case similarities [19].

LLM- and vision-language-based approaches, such as PathChat [24], Quilt-LLaVA [25], and HistoChat [26], are gaining popularity, but they do not solve the lack of explainable AI in pathology. Their strength is also their limitation: they are optimized to align images with text and generate likely diagnostic language, not to classify spatially distributed, multi-semantic morphologies through an explicitly inspectable representation. Image-text alignment makes these systems highly flexible, but it does not necessarily expose the internal features, representations, or reasoning mechanisms by which a diagnosis is reached. Instead, such models associate visual patterns with diagnostic text and generate the answer that best fits the learned image-text distribution. In clinical pathology, however, the highest-value output is not a single fluent answer, but a decision that supports the final diagnosis, identifies uncertainty, and helps rule in or rule out relevant differential diagnoses. Thus, text explanations generated by LLM-based systems can appear persuasive while still obscuring the model’s actual representation structure. Pathology therefore needs models that provide semantic explanations of internal model representations, linking predictions to morphologic concepts, cross-case similarities, marker patterns, and diagnostic differentials.

Concept learning can provide morphologically meaningful insights into the decision patterns of diagnostic AI. For example, supervised concept-learning approaches, such as concept bottleneck models, employ a predefined set of concepts and corresponding labels to generate insights about model recognition. However, these concepts are based on human-defined morphologic categories, which may not capture the ways in which visually similar regions are represented in the model’s latent feature space. Moreover, forcing predictions through a human-defined concept bottleneck can constrain the model to an incomplete semantic vocabulary, potentially reducing its predictive performance [27]. Human-defined concept spaces may therefore be insufficient for explaining the internal representations of foundation models. Unsupervised concept learning, in contrast, can identify recurring visual features as they are naturally represented by the model in latent space. At subsequent review and restriction to a pathologist-recognizable morphology, these model-derived concepts can provide a stronger basis for pathology-specific explanations, while also allowing non-morphologic or inconsistent concepts to be identified.

We built upon the CONCH v1.5 foundation model to develop an AI-based classifier for 19 sarcoma subtypes using a tissue microarray cohort of 2,545 cases represented by 195,952 image patches, trained in 3-fold cross validation. The resulting attention-based multiple instance learning (ABMIL) model achieved strong subtype prediction performance in a 3-fold cross-validation, with a balanced accuracy of 77.38% ±1.88 and an averaged one-vs-rest AUC of 0.98. We next implemented a interpretability pipeline based on a sparse autoencoder (SAE) and CONCH-derived embeddings to learn sparse, recurring visual concepts [28], filter and annotate them by three senior pathologists and then incorporate them into a linear, explainable diagnostic model. After training on the full SAE concept space, this linear model retained most of the original ABMIL performance, reaching 73.74% ± 1.3 balanced accuracy, and 67.04% ± 1.27 after training on pathologist-curated morphologic concepts only. Of note, the residual gain of the full SAE concept model was collated from a set of concepts that were either inconsistent, technical artifacts, or labelled as diagnostically irrelevant by pathologists, showing the performance gains from confounding, diagnostically irrelevant shortcuts rather than from clinically meaningful morphology.

Together, these findings show that CONCH v1.5 embeddings capture diagnostically relevant morphologic signal for sarcoma subtype classification, and that SAE-derived concepts can translate part of this signal into pathologist-reviewable features. More broadly, our results suggest that concept-based modeling can serve not only as an explanation tool, but as an foundation model auditing framework to distinguish performance supported by pathologist-recognizable morphology from performance driven by non-interpretable or non-diagnostic visual regularities.

## Methods

### Cohort and Dataset Creation

We assembled a development data set of 68 H&E-TMA slides, scanned using an Aperio AT2 scanner at 20x magnification (0.5013 mpp). Each TMA contained multiple TMA cores per patient, obtained from the same or different FFPE blocks. To extract individual core images, we segmented the tissue using HEST [29] via the TRIDENT [30] repository, converted all slide-level segmentations to GeoJSON rectangles, and imported them in QuPath [31]. We then reduced the annotations to one core per patient. We predefined inclusion criteria as a minimum of approximately 20% tissue retained per core and the absence of any obvious artifacts, such as dust particles, large out of focus regions, or apparent tissue folds. To avoid sampling bias, we applied a prespecified core-selection rule: the first available core was selected if it met inclusion criteria. If none of the cores per patient met the criteria, the patient was rejected from this study. In cases where cores overlapped, we drew a polygon around the core to be extracted; in all other cases, we used a rectangle as the bounding box. During core image extraction, we used these annotations uniformly to first obtain the final bounding box and then applied a binary mask to retain only the tissue within the annotation, setting all background pixels to white to avoid mixing tissue from adjacent cores. In total, we extracted cores from 2,545 patients spanning 19 sarcoma subtypes (Table 1).

**Table 1.** The subtypes, abbreviations, and TMA core count of our development dataset.

| Sarcoma Subtype | Abbreviation | Extracted Cores |
| --- | --- | --- |
| Angiosarcoma | AS | 142 |
| Chordoma | CDM | 130 |
| Clear Cell Sarcoma | CCS | 19 |
| De-differentiated Liposarcoma | DDLPS | 135 |
| Desmoplastic Small Round Cell Tumor | DSRCT | 315 |
| Epithelioid Sarcoma | EPS | 35 |
| Gastrointestinal Stromal Tumor | GIST | 128 |
| Malignant Peripheral Nerve Sheath Tumor | MPNST | 149 |
| Mesenchymal Chondrosarcoma | MC | 29 |
| Myxoid Liposarcoma | MLPS | 106 |
| Osteosarcoma | OS | 249 |
| PDGFRA-mutated Gastrointestinal Stromal Tumor | PDGFRAGIST | 12 |
| Pleomorphic Liposarcoma | PLS | 44 |
| Schwannoma | SW | 62 |
| Soft Tissue Leiomyosarcoma | STLMS | 205 |
| Synovial Sarcoma | SS | 170 |
| Undifferentiated Pleomorphic Sarcoma | UPS | 99 |
| Uterine Leiomyosarcoma | ULMS | 184 |
| Well-differentiated Liposarcoma | WDLPS | 332 |
| **Total count** |  | **2,545** |

We computed patch embeddings for each of the cores using CONCH 1.5 [32], an extended version of the CONCH histopathology vision-language foundation model [23], via TRIDENT on 256×256 patches at a 20x magnification, resulting in a total of 195,952 patches, stored at the core level.

### Computational Environment

All experiments were run on a local workstation equipped with a 32-core AMD Ryzen Threadripper PRO 5975WX CPU, an NVIDIA RTX A6000 GPU with 48 GB VRAM, and 512 GB system memory. The system ran Ubuntu 24.04.4 LTS with PyTorch 2.10.0+cu128. Model training and inference used the TRIDENT repository at commit bb45da57d2927b9e70bb4f1383c3455e428febb7 and the CONCH v1.5 model checkpoint 3e5766a5d1500d53c73c03005e24c30c1f27be13. This configuration was used to ensure reproducible feature extraction, model inference, and downstream analyses.

### AI Models

#### Sparse Autoencoder

We trained a sparse autoencoder (SAE) on patch-level, 768-dimensional CONCH 1.5 embeddings that had been extracted from the full development cohort. The SAE was trained once as an unsupervised concept-discovery step; it did not use sarcoma subtype labels, fold assignments, or downstream classification outcomes. The SAE pipeline was implemented in Python using PyTorch, with additional libraries for data handling and orchestration. Core-level patches were treated as feature bags defined in a study-wide manifest file. We used a one-layer encoder and one-layer decoder architecture with a tied bias term initialized from the geometric median of the embeddings. The expansion factor was set to 1, yielding 768 learned SAE features. The model was trained to minimize reconstruction loss with an L1 sparsity penalty on the learned activations. Training was performed using the Adam optimizer at a learning rate of 1e-4, using mean squared error averaged over embedding dimensions as reconstruction loss, and the mean L1 norm of the activation vector as a sparsity penalty, with an L1 coefficient of 0.004. After training, the SAE encoder was fixed and used to project each patch embedding into concept space, preserving coordinates and available metadata for downstream classification and interpretation. Supervised classification models were subsequently trained and evaluated within the cross-validation framework using these fixed SAE-derived concept representations.

#### Concept Annotations

For concept annotation, concepts were ranked by mean activation across all patches in the development cohort. The concept review set was defined by the sharp inflection point in the ranked activation distribution (Figure 2B), after which mean activation dropped to near-background levels. This cutoff corresponded to a mean activation of 0.001 and resulted in 90 SAE-derived concepts. For each selected concept, we generated a review sheet containing 16 representative patches selected by maximum concept activation. The reviewing pathologists were provided with the corresponding sarcoma subtype labels and concept activation scores. Each concept was annotated using four fields: free-text concept name, subtype relevance among the 19 included sarcoma subtypes, binary usability rating (usable Y/N), and optional notes. Disagreement in usability ratings was resolved in a subsequent consensus meeting. Interobserver agreement was summarized as percent agreement.

#### ABMIL Model

For the attention-based multiple instance learning (ABMIL) model, each TMA core was treated as a bag of CONCH v1.5 patch embeddings, with the core label corresponding to the sarcoma subtype. Model training and evaluation were performed in a 3-fold cross-validation at the patient/core level with subtype-stratified folds. Within each run, final checkpoint selection was performed on the validation fold; the held-out test fold was used only for final performance estimation. We trained a class-wise gated ABMIL model, optimized for up to 300 epochs, using Adam with a learning rate of 5×10^−5^ and a weight decay of 5×10^−6^ (with balanced softmax loss and early stopping, based on validation balanced accuracy with a patience of 15 epochs). The final checkpoint was selected based on the highest validation balanced accuracy. After checkpoint selection, we applied post-hoc logit adjustment by adding class-specific bias terms to the logits. These bias terms were adjusted on the validation set over 10 rounds to maximize validation balanced accuracy, using candidate values from −0.5 to 0.5 in increments of 0.1.

To explore the effect of sample size on model performance, we performed a case-per-class scaling experiment, using the same 3-fold cross-validation splits and the same ABMIL architecture, training procedure, and hyperparameters described above. Within each fold, cases were randomly subsampled within the training, validation, and test sets, while preserving the original split assignment. Rare classes were retained in the analysis and included up to the maximum number of available cases. For each sampled case-per-class setting, we trained the ABMIL model once and evaluated class-wise accuracy, defined as the proportion of correctly classified cases within each true class, and class-wise AUC. Balanced accuracy was calculated as the unweighted mean of class-wise accuracy across all included classes.

#### ProtoMIL Model

We trained a linear ProtoMIL model [28] on SAE–derived concept activations, filtered by expert-curated annotations. This linear model provides weighted concept contributions, enabling case-level interpretability. We allowed the model to train for 100 epochs using Adam (learning rate 1×10^−4^, weight decay of 1×10^−4^), optimized on balanced softmax loss, and then selected the final model checkpoint, based on the highest balanced accuracy on the validation set, by using an early stopping criterion (patience 10).

For case-level explanations, we selected the top 4 contributing concepts per core, based on the summed positive signed contribution across all patches in the bag, ignoring negative contribution. For each selected concept, patch-level contributions were mapped back to their spatial coordinates to generate heatmaps, which were normalized to the maximum activation across the four selected concepts within the same core.

## Results

### CONCH-based MIL captures subtype-discriminative morphology in sarcoma

To assess whether CONCH v1.5 embeddings capture subtype-discriminative signal, we trained a gated class-wise ABMIL model in a 3-fold cross-validation with balanced softmax and posthoc-logit adjustment (Figure 1A). We then selected the checkpoint with the highest balanced accuracy on the validation dataset, achieving a mean balanced accuracy of 77.38% (± 1.88) and mean AUC of 97.85% (± 0.4) across the test sets (Figure 1B). Class-wise performance for each specific diagnosis was heterogeneous (sensitivity of 59.2–94.7%). The model prediction for CCS, EPS, and CDM showed high sensitivity (>90%), with minimal confusion to other diagnoses. SS, SW, MLPS, PDGFRAGIST, and GIST also performed strongly (≈81–88%), with limited spread into related classes.

**Figure 1.**
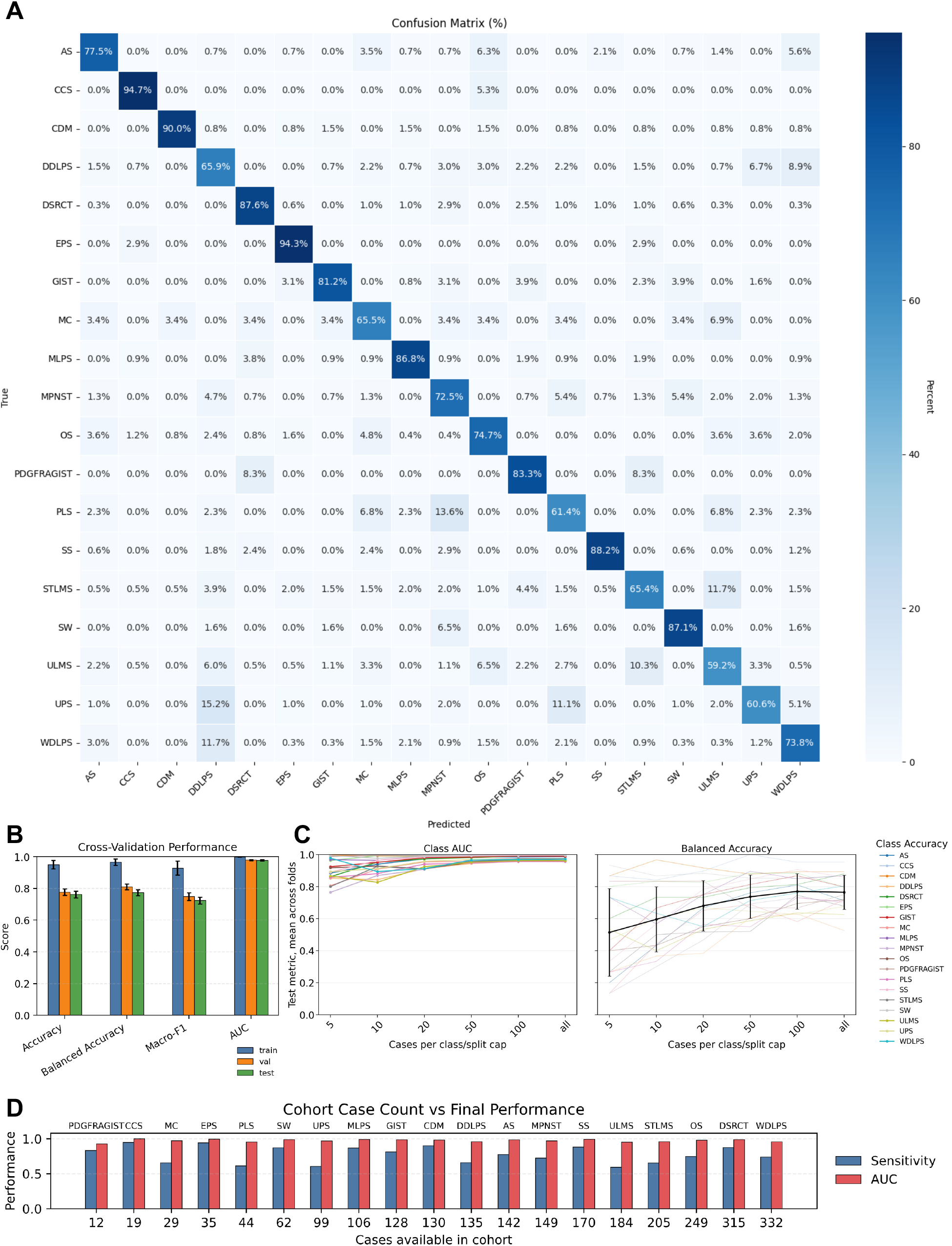
Model performance across sarcoma subtypes and data scaling. (A) Confusion matrix showing class-wise sensitivity across 19 sarcoma subtypes. (B) Cross-validation performance across train, validation, and test splits, showing accuracy, balanced accuracy, macro-F1, and macro-averaged AUC. (C) Case-per-class scaling analysis: class-wise AUC saturates early, while balanced accuracy improves with increasing sample size. (D) Relationship between cohort size and final performance: AUC remains high across classes, whereas sensitivity does not show a simple relationship with class size.

Not surprisingly, most of the errors in prediting the correct diagnosis were concentrated in morphologically related groups. Liposarcoma subtypes showed bidirectional confusion: DDLPS was frequently misclassified as WDLPS (8.9%), and vice versa (11.7%), with additional spillover into UPS (6.7%). Smooth muscle tumors also showed overlap: ULMS and STLMS were commonly confused (10.3% and 11.7%, respectively). In addition, ULMS was confused with OS (6.5%), while STLMS was confused with both PDGFRA-mutated GIST (4.4%) and DDLPS (3.9%). The pleomorphic entities (PLS, UPS, MPNST) formed a broad confusion cluster (ranging from 2.0% to 13.6%). Errors for AS were dispersed into multiple classes, rather than any single dominant confusion.

GIST and PDGFRA-mutant GIST remained relatively well separated, but GIST showed minor confusion with its PDGFRA-mutated phenotype (3.6%). MC showed moderate performance with scattered errors, with the highest confusion seen for ULMS (6.9%). DSRCT retained high sensitivity (87.6%), but with low-level dispersion into other subtypes (up to 2.9% with MPNST).

In a case-per-class scaling experiment using fixed cross-validation splits, performance appeared to show diminishing gains with sample sizes of approximately 100 cases per class (Figure 1C). In addition, class-wise AUC reached near-saturation before this point; balanced accuracy, however, continued to improve up to 100 cases before showing smaller additional gains with reduced variance across folds. Beyond 100 cases per diagnosis, additional data produced minimal gains.

In our cohort, final performance was not simply determined by class size (Figure 1D): several low-count classes achieved high accuracy, whereas multiple higher-count classes remained comparatively lower. By contrast, AUC was high across classes and showed no clear relationship with sample count.

To contextualize the difficulty of the 19-class sarcoma classification task, we summarized published diagnostic concordance studies in soft tissue sarcoma pathology. Since subtype-specific sensitivity based on H&E alone is rarely reported, we used diagnostic agreement studies that compared initial diagnoses with expert or centralized review as a proxy (Table 2).

**Table 2.** Published diagnostic agreement values across human pathologists in sarcoma pathology, based on H&E and ancillary-test supported diagnostic workflows.

| Agreement | Major Disagreement | Minor Disagreement | Year | Reference |
| --- | --- | --- | --- | --- |
| 54% | 19% | 27% | 2009 | [33] |
| 73.4% | 10.9% | 15.6% | 2010 | [34] |
| 71.8% | 16.4% | 11.8% | 2014 | [3] |
| 71% | 8% | 21% | 2016 | [35] |
| 86% | – | – | 2018 | [36] |
| 64.2% | 24.3% | 11.5% | 2024 | [37] |
| 44% | 44% | 12% | 2025 | [38] |

Across published studies, diagnostic concordance in soft tissue sarcoma has varied widely, with overall agreement ranging from 44% to 86%, while major disagreement rates range from 8% to 44%. Importantly, these agreement values generally reflect clinical diagnostic workflows that may include H&E morphology together with immunohistochemistry, molecular testing, clinical information, and expert consultation. These rates, however, are not directly comparable with those of our model, which was trained and evaluated solely using H&E morphology from TMA cores.

Nevertheless, these agreement values provide useful clinical context for the difficulty of sarcoma subtype classification. In our H&E-only TMA-core setting, the ABMIL model achieved a balanced accuracy of 77.38% ± 1.88 and a macro-averaged AUC of 97.85% ± 0.4 in 19-class cross-validation, demonstrating that foundation-model embeddings capture substantial morphologic signal for sarcoma subtype prediction on H&E.

### Unsupervised concept learning can identify morphologic features distinctive for specific sarcoma subtypes

To obtain morphologically meaningful explanations, we investigated whether concept learning could identify recurrent image patterns associated with specific sarcoma subtypes. In supervised concept learning, pathologists would need to pre-define and annotate morphologic features across large numbers of image patches from different diagnostic classes. In contrast to this labor-intensive approach, our unsupervised workflow used an SAE to transform CONCH-derived patch embeddings into a finite concept space, shifting pathologist review from individual patches to recurring model-derived image patterns (Figure 2A). We configured our SAE with an expansion factor of 1, yielding 768 concepts in total. From a total of 2,545 core images, we extracted 195,952 patches. We then measured the mean activation per concept across all patches in the dataset and determined 0.001 mean activation as the cutoff, resulting in 90 concepts (Figure 2B). Next, we engaged three board-certified senior pathologists (AJI, YPH, AJL) to individually rate the obtained concepts, describe their associations to distinct sarcoma subtypes, if applicable, and rate their usability in AI predictions. In a subsequent consensus meeting, we agreed on a final set of concept labels.

**Figure 2.**
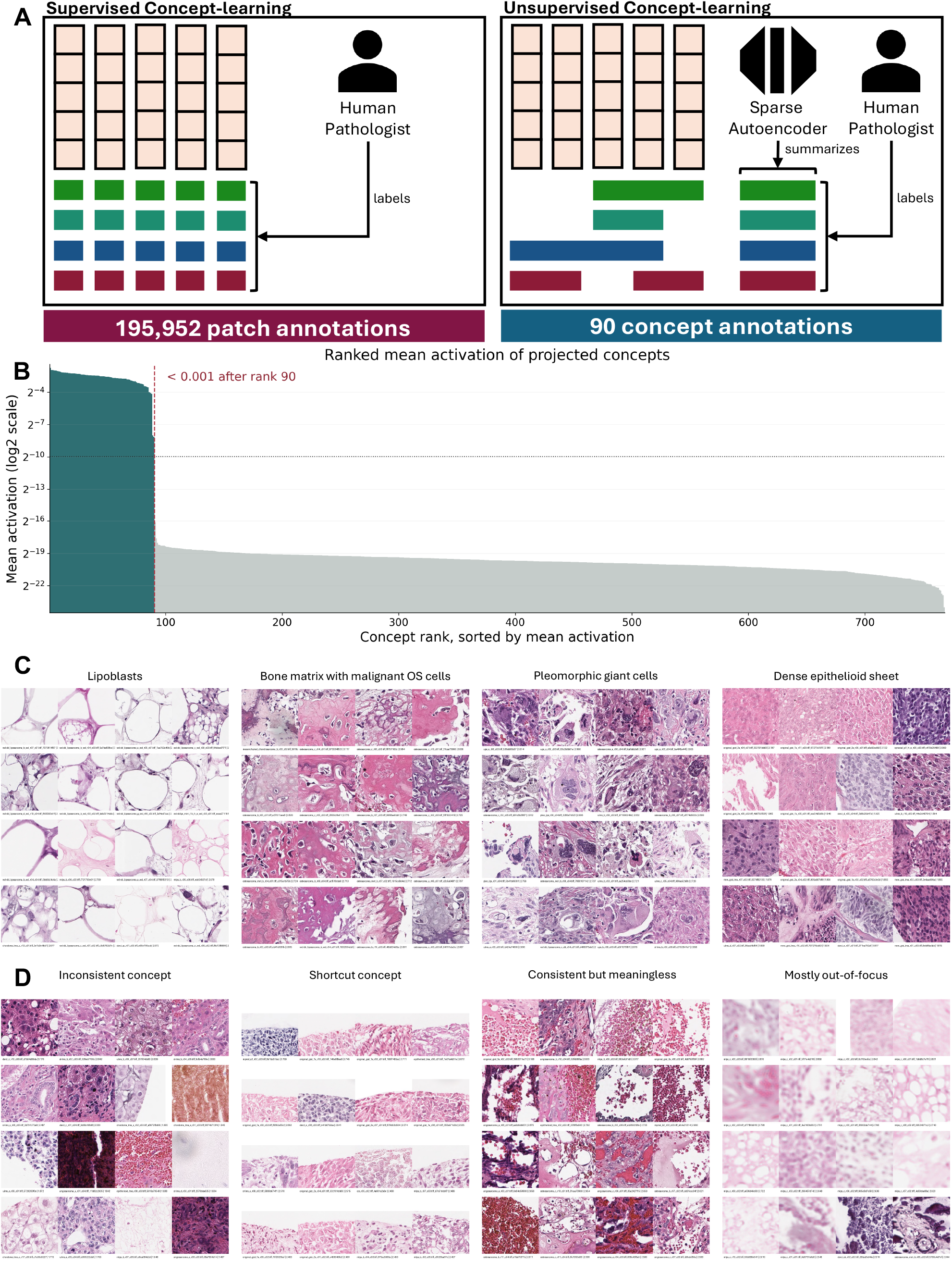
Unsupervised concept learning and expert annotations. (A) Comparison of required annotations for supervised concept learning and unsupervised concept learning. (B) Ranked mean activation of concepts, capped below an activation of 0.001. (C) Examples of high-activation concepts corresponding to recognizable histomorphological patterns. (D) Examples of high-activation concepts capturing structural, histopathologically irrelevant patterns.

Interpretation of concept relevance showed substantial variation between pathologists. From the 90 reviewed concepts, the three pathologists agreed on usability in 58 concepts (27 useful 31 not useful) during independent concept review. Of the remaining 32 concepts, they agreed that 14 were useful and 18 were not useful. Following a consensus meeting, we agreed on a final set of 41 useful concepts (Figure 2C). Among the concepts deemed not useful, we recognized three distinct categories: (1) inconsistent concepts, (2) consistent patches that aren’t morphologically meaningful, and (3) scanning artifacts (Figure 2D). We found 23 inconsistent concepts, in which some patches seemed to follow a shared feature, while others clearly didn’t (e.g. intact vs fragmented tissue in combination with clear cells, physaliferous cells, and spindle cells). In 25 concepts, we could identify consistent but meaningless patterns, such as a prominent red dot in each of the patches, crowded red blood cells, similarly distributed tissue fragments, or tissue in only one half or corner of the image. Finally, we found one concept that focused mostly on out-of-focus patches.

### ProtoMIL models provide explainability in predicting sarcoma subtypes from concept activation vectors

We next compared raw CONCH-based ABMIL with concept-based model variants to assess the tradeoff between predictive performance and interpretability. Raw ABMIL trained directly on CONCH embeddings achieved the highest balanced accuracy of 77.38% ±1.88. Among concept-based models, linear ProtoMIL trained on the full 768-dimensional SAE concept space retained much of the raw ABMIL performance (73.74% ± 1.3) while providing concept-level interpretability. Restricting ProtoMIL to pathologist-curated morphologic concepts further reduced balanced accuracy from 73.74% ± 1.30% to 67.04% ± 1.27%. ABMIL, however, when trained on the concept vector, performed worse compared to ProtoMIL, achieving 67.05% ± 1.51 on the full concept vector and 61.01% ± 2.31 on the meaningful concepts. Thus, within the SAE concept space, a substantial fraction of subtype-discriminative signal was retained in expert-interpretable concepts, while the residual performance gains when trained on the full concept vector was attributable to information carried by non-curated concepts, including technical artifacts, inconsistent concepts, and diagnostically irrelevant visual similarities.

Concept explanations summarize the relative contribution of individual concepts and can be linked back to attention maps. While the fractions of the top contributing concepts already provide an interpretable summary, mapping these concepts back to the tissue further clarifies how the model combines morphologic evidence. In a case of pleomorphic liposarcoma, for example, the patch with the strongest contribution to the classification did not show the highest activation of any single concept. Instead, it combined several moderately activated concepts, including pleomorphic giant cells, lipoblasts, and atypical spindle cells, within the same patch (Figure 3B). The heatmap highlighted two coherent tumor regions and a few additional individual patches, all showing characteristic features of pleomorphic liposarcoma. This suggests that the model did not simply select patches with the strongest single-concept activation, but favored regions in which multiple diagnostic concepts co-occurred. Such information would not be available from conventional attention heatmaps alone.

**Figure 3.**
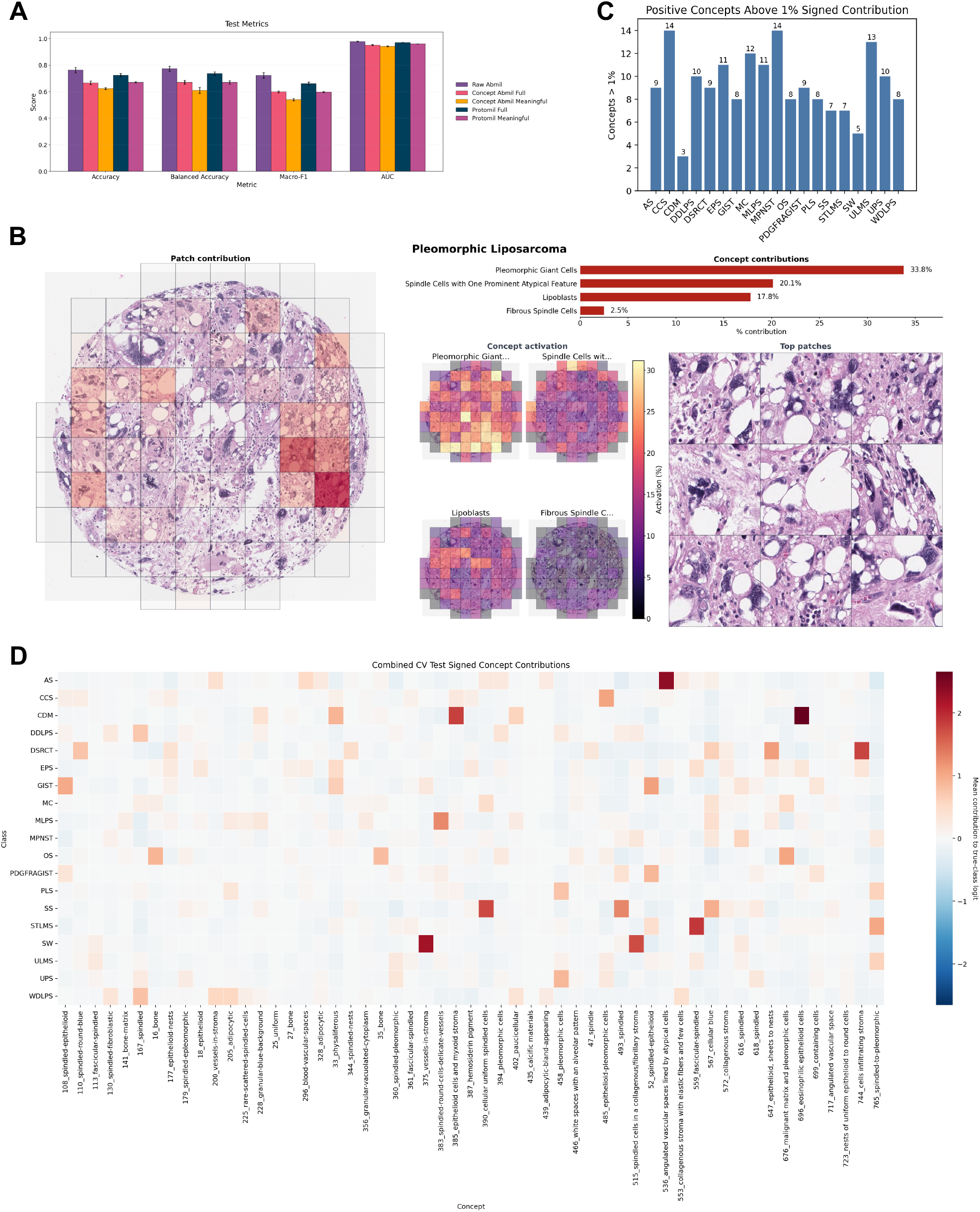
Concept-based interpretability of subtype predictions. (A) Comparison of model variants, showing performance across accuracy, balanced accuracy, macro-F1, and one-vs-rest AUC for ABMIL on the raw embeddings, full concept vector, and meaningful concepts, as well as ProtoMIL on the full concept vector, and on meaningful concepts. (B) Example case (Pleomorphic Liposarcoma) showing slide-level explanations for the top contributing concepts, alongside spatial heatmaps highlighting concept localization within the tissue. (C) Histogram showing the number of concepts that positively contribue by at least 1% to model decisions by the ProtoMIL model on meaningful concepts. (D) Heatmap of signed concept contributions aggregated across cross-validation folds, illustrating ProtoMIL’s representation of subtype-associated morphology.

#### Model Explanations by Subtype

Concept-based explanations of sarcoma subtype predictions aligned with known morphological features. For each subtype, we identified concepts with at least 1% positive contribution, yielding 3 to 14 contributing concepts per subtype (Figure 3C). We further examined the mean activation of these concepts within each subtype, providing an overview of the model’s representation of subtype-associated morphology (Figure 3D). Together, these analyses showed that the concept-based explanations reflected sarcoma-specific morphologic patterns.

Liposarcoma subtypes showed the most subtype-specific profiles: MLPS was dominated by small round blue cells in a myxoid matrix, whereas PLS was characterized by pleomorphic giant cells, lipoblasts, and atypical spindle-cell concepts. WDLPS and DDLPS shared fibrous spindle-cell and collagenous stromal concepts, while DDLPS showed little contribution from adipocytic concepts.

High-grade pleomorphic tumors showed clear separation. OS was explained predominantly by malignant bone matrix and laminar bone concepts, whereas UPS was explained by pleomorphic giant cells, mitotic pleomorphic spindle cells, dense spindle-cell sheets, and poorly differentiated spindle-cell concepts.

Smooth muscle tumors were primarily explained by atypical and eosinophilic fascicular spindle-cell concepts. STLMS showed stronger contribution from a dense eosinophilic fascicular-spindled concept, whereas ULMS showed greater contribution from mitotic and pleomorphic spindle-cell concepts.

Among peripheral nerve sheath tumors, schwannoma was enriched for stromal vascular and collagenous spindle-cell concepts, whereas MPNST was enriched for variable fascicular spindle-cell, atypical spindle-cell, and dense spindle-cell concepts. Of note, the physaliferous concept was part of the MPNST-explanation, which points to label-ambiguity and geometric similarity to vacuolated cytoplasm rather than a truly physaliferous component in MPNST.

Small round cell sarcomas showed greater concept overlap. DSRCT was distinguished by infiltrating epithelioid nests, dense round blue cell sheets, and fibrous matrix, whereas SS was dominated by dense spindle-cell fascicular concepts. MC showed enrichment for matrix-associated concepts, including malignant bone matrix and laminar bone.

## Discussion

Our results demonstrate that foundation-model-derived H&E features contain strong morphologic signal relevant to the challenging task of sarcoma subtyping, but also underscore the current limitations of H&E-only AI for clinical-grade diagnosis. Using CONCH v1.5 embeddings, the gated class-wise ABMIL model achieved high cross-validated performance across 19 sarcoma subtypes, with a macro-averaged AUC of 97.85% and a balanced accuracy of 77.38% ±1.88. This performance is notable for two reasons. First, sarcoma morphology overlaps substantially across subtypes, and published diagnostic agreement rates from 44% to 86% [33, 34, 3, 35, 36, 37, 38] reflect the difficulty of this diagnostic space, even in full clinical workflows that include ancillary testing beyond H&E alone. Second, the high macro-averaged AUC shows that the model performs well across individual diagnostic classes, rather than just doing well on average. This matters in pathology because a model that can predict the set of most likely diagnoses is often more useful to a pathologist than a black-box model that simply outputs a single diagnosis [39]. However, the balanced accuracy of 77.38% indicates that even a rigorously trained model using state-of-the-art foundation-model embeddings is not sufficient as a stand-alone clinical diagnostic tool. This limitation is especially important because our study focused on only a subset of sarcoma subtypes and did not include the full spectrum of sarcoma mimickers encountered in routine practice, making the classification task less complex than real-world sarcoma diagnosis. Together, these findings support the feasibility of explainable, morphology-based AI assistance in sarcoma classification on H&E, while emphasizing the need for multimodal diagnostic frameworks that incorporate immunophenotypic, molecular, and contextual information.

The observed subtype confusions were largely consistent with known diagnostic difficulties in sarcoma pathology: liposarcomas separated according to expected adipocytic, myxoid, fibrous, and pleomorphic features, with overlap reflecting recognized morphologic continua [40, 41, 42, 43, 44]. GIST and PDGFRA-GIST shared spindle/epithelioid concepts, consistent with their known morphologic spectrum, although PDGFRA-associated features were only partially captured [15, 17]. Smooth-muscle tumors clustered around fascicular spindle and spindled-pleomorphic concepts, in line with leiomyosarcoma morphology, but without specific smooth-muscle cytologic features [45, 46]. Schwannoma and MPNST shared spindle/fibrillary stromal concepts, reflecting their peripheral nerve sheath differentiation, while MPNST showed more atypical spindle-cell activations [47, 48]. Small round blue cell tumors showed substantial overlap, with DSRCT, mesenchymal chondrosarcoma, and synovial sarcoma separated mainly by epithelial-nested, matrix-forming, or dense spindle-cell concepts [49, 50, 51]. Epithelioid tumors were distinguished by vascular concepts in angiosarcoma, epithelioid/myxoid features in CCS, and physaliferous or infiltrative epithelioid concepts in EPS, although in everyday diagnostics, vascular concepts may be equally associated with benign vascular mimics such as anastomosing hemangioma [52, 53, 54, 55]. Finally, osteosarcoma and UPS separated clearly through bone-forming versus pleomorphic spindle-cell concepts, consistent with their defining histology and the diagnosis-of-exclusion nature of UPS [56, 57].

SAEs have been used in digital pathology before, mainly for tasks like stain normalization and nuclei detection [58, 59]. More recent studies have shown that SAEs can help understand which biological representations pathology foundation models have learned [60]. Our study builds most directly on ProtoMIL, which uses SAE-derived concepts from CONCH embeddings to build an interpretable WSI-classification model. Instead of relying only on opaque slide-level predictions, ProtoMIL represents cases through human-inspectable concepts that can be labelled, curated, and filtered to remove spurious signals [28]. More broadly, concept-based interpretability is becoming an important direction in computational pathology, with prior work showing that interpretable image features can capture tissue composition and architecture using sparse group lasso models [61], and that hierarchical concepts can be linked to foundation-model predictions using Shapley attribution [62]. Building on this work, a central contribution of our study is applying SAE-derived concepts to sarcoma pathology, where we produced a finite set of recurring visual concepts that could be reviewed and annotated by pathologists. This shifts interpretation from isolated image localization toward concept-level explanation, asking which visual patterns the model represents, which concepts contributed to a prediction, and whether those concepts correspond to recognizable histopathology.

AI models can learn spurious signal, dataset-specific distributions, and noise that artificially inflate performance [63, 64, 65]. Restricting the model to meaningful concepts only provides a direct way to test how much performance was supported by pathologist-recognizable signal, and how much was gained from concepts that pathologists would not consider diagnostically meaningful. Raw ABMIL remained the strongest classifier, indicating that unconstrained CONCH embeddings retain predictive signal not fully captured by the SAE concept models. However, ProtoMIL trained on the full SAE concept activation vector retained much of the original ABMIL performance while providing concept-level interpretability. When ProtoMIL was restricted to pathologist-curated morphologic concepts, mean balanced accuracy decreased from 73.74% to 67.04%, indicating that most subtype-discriminative signal within the concept space remained available in expert-interpretable concepts. The residual gain of the full concept model came from non-curated concepts, including technical artifacts, spurious patterns, and diagnostically irrelevant visual similarities. Thus, higher performance within the unconstrained concept space did not necessarily reflect better diagnostic evidence. SAE-derived concept modeling therefore provides a way to estimate the morphology-supported fraction of concept-based diagnostic performance.

Several limitations define the scope of this study. The cohort consisted of TMA cores rather than whole-slide diagnostic specimens, limiting assessment of intratumoral heterogeneity, necrosis, dedifferentiation, and spatial transitions between tumor components. Furthermore, we considered only a single magnification level. Pathologists operate on different magnification levels in their daily work, and an investigation of concepts on other scales as well as multi-scale concepts should be further investigated. The focus on individual high-resolution patches may also explain the lower performance for some sarcoma types whose diagnosis depends on the combined presence of multiple features, such as mesenchymal chondrosarcoma, which is biphasic by definition. Although ABMIL can theoretically model relationships between regions, the data and labeled concepts must provide enough evidence for the model to represent them. Finally, as we trained the SAE on the full cohort used for model development, it remains to be investigated how well SAE-derived concepts can explain models trained on an independent dataset.

All results are based on internal cross-validation and require external validation across institutions, scanners, staining protocols, and routine biopsy or resection material. Some subtypes were represented by few cases, limiting precision of class-specific estimates. Furthermore, the model intentionally used H&E morphology alone; clinical sarcoma diagnosis still requires integration with IHC, molecular findings, anatomic site, and clinical context. Accordingly, the current model should be viewed as a morphology-based decision-support and interpretability framework, not as a stand-alone diagnostic system. Finally, our dataset did not include any control tissue, abstracting away non-sarcomatoid differential diagnoses and benign tissue.

Future work should test whether this framework generalizes to biopsies and whole-slide clinical material, and whether SAE-derived concepts remain stable across independent cohorts. The high one-versus-rest discrimination suggests a natural next step: using ranked H&E-based predictions to prioritize ancillary testing, such as IHC or molecular assays that best resolve the leading differential diagnoses. Spatial mapping of concept activations may also enable concept-level analysis of heterogeneous tumor regions. More broadly, SAE-derived concepts could provide a structured morphologic interface between foundation models, pathologists, and diagnostic support systems. The recurring categories we observed within the concept labels, including cell phenotype, cytologic appearance, extracellular matrix characterization, tissue architecture, degree of organization, and density, may form the basis for structured concept-annotation frameworks.

## Conclusion

In summary, this study shows that H&E morphology contains substantial subtype-discriminative signal across a large and heterogeneous sarcoma cohort. Using CONCH-derived features from 2,545 cases spanning 19 entities, we found that several sarcoma subtypes can be separated from H&E alone with high one-versus-rest performance, while single-label classification remains limited by recognized morphologic overlap between related diagnoses. Importantly, SAE-derived concepts recovered recurring tissue concepts that expert pathologists recognized as characteristic for many sarcoma subtypes. These concepts reflected known diagnostic patterns, including lipogenic, myxoid, spindle-cell, pleomorphic, vascular, small round blue cell, and matrix-forming morphologies, while less prevalent but diagnostically important patterns remained incompletely captured. Finally, we demonstrated that a non-trivial fraction of model performance can be attributed to diagnostically irrelevant concepts, including inconsistent concepts and technical artifacts, showing that predictive performance alone can overestimate diagnostically meaningful performance. Together, these findings show what can already be extracted from H&E in rare sarcoma pathology and where H&E-based AI reaches its current limits. With larger and more diverse cohorts, this approach may allow increasingly granular morphologic concept spaces to be learned, curated, and linked to diagnostic reasoning across rare tumor entities. More broadly, our results suggest that concept-based modeling can move foundation-model interpretability beyond attention maps toward an auditable morphologic vocabulary, providing a framework for studying, explaining, and extending AI-assisted diagnostic reasoning in rare, heterogeneous tumors.

## Data Availability

Raw H&E-stained whole-slide images and associated case-level metadata are not publicly available. Source code, concept dictionaries, and trained model weights will be made publicly available upon peer-reviewed publication of the manuscript. Requests for access to additional materials may be directed to the corresponding author and will be considered subject to applicable approvals.

## Statements

### Ethics Approval

The Institutional Review Board of Mass General Brigham gave ethical approval for this work under protocol number 2025P000516.

## References

[1] Sbaraglia M, Bellan E, Dei Tos AP. The 2020 WHO Classification of Soft Tissue Tumours: news and perspectives. Pathologica 2021;113:70–84.

[2] Burningham Z, Hashibe M, Spector L, Schiffman JD. The epidemiology of sarcoma. Clin Sarcoma Res 2012;2:14.

[3] Thway K, Wang J, Mubako T, Fisher C. Histopathological diagnostic discrepancies in soft tissue tumours referred to a specialist centre: reassessment in the era of ancillary molecular diagnosis. Sarcoma 2014;2014:686902.

[4] Wilson R, Reinke D, van Oortmerssen G, Gonzato O, Ott G, Raut CP, et al. What is a sarcoma ‘specialist center’? Multidisciplinary research finds an answer. Cancers (Basel) 2024;16:1857.

[5] Wang, X. Q., Wang, X. Q., Hsu, A. T. Y. W., Goytain, A., Ng, T. L. T., & Nielsen, T. O. (2021). A Rapid and Cost-Effective Gene Expression Assay for the Diagnosis of Well-Differentiated and Dedifferentiated Liposarcomas. The Journal of molecular diagnostics: JMD, 23(3), 274–284. 10.1016/j.jmoldx.2020.11.011

[6] Vats, K., Spafford, M., Groot, G., Graham, P., Banerjee, T., Deobald, R., & Osmond, A. (2022). Moving towards the optimization of diagnosis for patients with sarcoma: A 10-year review of externally consulted sarcoma cases in a general anatomical pathology service. Annals of diagnostic pathology, 60, 151958. 10.1016/j.anndiagpath.2022.151958

[7] Choi JH, Ro JY. The recent advances in molecular diagnosis of soft tissue tumors. Int J Mol Sci 2023;24.: 10.3390/ijms24065934.

[8] Pestana RC, Lopes David BB, Pires de Camargo V, Munhoz RR, Lopes de Mello CA, González Donna ML, et al. Challenges and opportunities for sarcoma care and research in Latin America: a position paper from the LACOG sarcoma group. Lancet Reg Health Am 2024;30:100671.

[9] Dey, B., Srinivas, B. H., Badhe, B., Nachiappa Ganesh, R., Gochhait, D., Toi, P. C., & Jinkala, S. (2020). Malignant Epithelioid Soft Tissue Tumours-A Pathologist’s Perspective With Review of Literature. Cureus, 12(12), e12263. 10.7759/cureus.12263

[10] Wei, H., Zhen, T., Tuo, Y., Li, H., Liang, J., Chen, S., Shi, H., & Han, A. (2022). Clinicopathologic and molecular features of vascular tumors in a series of 118 cases. American journal of translational research, 14(5), 2939–2951.

[11] Zhang, L., Wang, R., Xu, R., Qin, G., & Yang, L. (2018). Extraskeletal Myxoid Chondrosarcoma: A Comparative Study of Imaging and Pathology. BioMed research international, 2018, 9684268. 10.1155/2018/9684268

[12] Mack, T., & Purgina, B. (2022). Updates in Pathology for Retroperitoneal Soft Tissue Sarcoma. Current oncology (Toronto, Ont.), 29(9), 6400–6418. 10.3390/curroncol29090504

[13] Thway K. (2019). Well-differentiated liposarcoma and dedifferentiated liposarcoma: An updated review. Seminars in diagnostic pathology, 36(2), 112–121. 10.1053/j.semdp.2019.02.006

[14] Orvieto, E., Furlanetto, A., Laurino, L., & Dei Tos, A. P. (2001). Myxoid and round cell liposarcoma: a spectrum of myxoid adipocytic neoplasia. Seminars in diagnostic pathology, 18(4), 267–273.

[15] Miettinen, M., & Lasota, J. (2011). Histopathology of gastrointestinal stromal tumor. Journal of surgical oncology, 104(8), 865–873. 10.1002/jso.21945

[16] Kobayashi, M., Inaguma, S., Raffeld, M., Kato, H., Suzuki, S., Wakasugi, T., Mitsui, A., Kuwabara, Y., Lasota, J., Ikeda, H., Miettinen, M., & Takahashi, S. (2019). Epithelioid variant of gastrointestinal stromal tumor harboring PDGFRA mutation and MLH1 gene alteration: A case report. Pathology international, 69(9), 541–546. 10.1111/pin.12830

[17] Daum, O., Grossmann, P., Vanecek, T., Sima, R., Mukensnabl, P., & Michal, M. (2007). Diagnostic morphological features of PDGFRA-mutated gastrointestinal stromal tumors: molecular genetic and histologic analysis of 60 cases of gastric gastrointestinal stromal tumors. Annals of diagnostic pathology, 11(1), 27–33. 10.1016/j.anndiagpath.2006.10.002

[18] Shaktah, L. A., Carrero, Z. I., Hewitt, K. J., Gustav, M., Cecchini, M., Foersch, S., Berezowska, S., & Kather, J. N. (2025). Application of artificial intelligence and digital tools in cancer pathology. The Lancet. Digital health, 7(10), 100933. 10.1016/j.landig.2025.100933

[19] Klauschen, F., Dippel, J., Keyl, P., Jurmeister, P., Bockmayr, M., Mock, A., Buchstab, O., Alber, M., Ruff, L., Montavon, G., & Müller, K. R. (2024). Toward Explainable Artificial Intelligence for Precision Pathology. Annual review of pathology, 19, 541–570. 10.1146/annurev-pathmechdis-051222-113147

[20] Chen, R. J., Ding, T., Lu, M. Y., Williamson, D. F. K., Jaume, G., Song, A. H., Chen, B., Zhang, A., Shao, D., Shaban, M., Williams, M., Oldenburg, L., Weishaupt, L. L., Wang, J. J., Vaidya, A., Le, L. P., Gerber, G., Sahai, S., Williams, W., & Mahmood, F. (2024). Towards a general-purpose foundation model for computational pathology. Nature medicine, 30(3), 850–862. 10.1038/s41591-024-02857-3

[21] Vorontsov, E., Bozkurt, A., Casson, A., Shaikovski, G., Zelechowski, M., Severson, K., Zimmermann, E., Hall, J., Tenenholtz, N., Fusi, N., Yang, E., Mathieu, P., van Eck, A., Lee, D., Viret, J., Robert, E., Wang, Y. K., Kunz, J. D., Lee, M. C. H., Bernhard, J. H.,… Fuchs, T. J. (2024). A foundation model for clinical-grade computational pathology and rare cancers detection. Nature medicine, 30(10), 2924–2935. 10.1038/s41591-024-03141-0

[22] Xu, H., Usuyama, N., Bagga, J., Zhang, S., Rao, R., Naumann, T., Wong, C., Gero, Z., González, J., Gu, Y., Xu, Y., Wei, M., Wang, W., Ma, S., Wei, F., Yang, J., Li, C., Gao, J., Rosemon, J., Bower, T.,… Poon, H. (2024). A whole-slide foundation model for digital pathology from real-world data. Nature, 630(8015), 181–188. 10.1038/s41586-024-07441-w

[23] Lu, M. Y., Chen, B., Williamson, D. F. K., Chen, R. J., Liang, I., Ding, T., Jaume, G., Odintsov, I., Le, L. P., Gerber, G., Parwani, A. V., Zhang, A., & Mahmood, F. (2024). A visual-language foundation model for computational pathology. Nature medicine, 30(3), 863–874. 10.1038/s41591-024-02856-4

[24] Lu, M. Y., Chen, B., Williamson, D. F. K., Chen, R. J., Zhao, M., Chow, A. K., Ikemura, K., Kim, A., Pouli, D., Patel, A., Soliman, A., Chen, C., Ding, T., Wang, J. J., Gerber, G., Liang, I., Le, L. P., Parwani, A. V., Weishaupt, L. L., & Mahmood, F. (2024). A multimodal generative AI copilot for human pathology. Nature, 634(8033), 466–473. 10.1038/s41586-024-07618-3

[25] Seyfioglu, M. S., Ikezogwo, W. O., Ghezloo, F., Krishna, R., & Shapiro, L. (2024). Quilt-llava: Visual instruction tuning by extracting localized narratives from open-source histopathology videos. In Proceedings of the IEEE/CVF Conference on Computer Vision and Pattern Recognition (pp. 13183–13192).

[26] Afzaal, U., Su, Z., Sajjad, U., Stack, T., Lu, H., Niu, S., Akbar, A. R., Gurcan, M. N., & Niazi, M. K. K. (2025). HistoChat: Instruction-tuning multimodal vision language assistant for colorectal histopathology on limited data. Patterns (New York, N.Y.), 6(8), 101284. 10.1016/j.patter.2025.101284

[27] Havasi, M., Parbhoo, S., & Doshi-Velez, F. (2022). Addressing leakage in concept bottleneck models. Advances in Neural Information Processing Systems, 35, 23386–23397.

[28] Sun, S., van Midden, D., Litjens, G., Baumgartner, C.F. (2026). Prototype-Based Multiple Instance Learning for Gigapixel Whole Slide Image Classification. In: Gee, J.C., et al. Medical Image Computing and Computer Assisted Intervention – MICCAI 2025. MICCAI 2025. Lecture Notes in Computer Science, vol 15973. Springer, Cham. 10.1007/978-3-032-05185-1_49

[29] Jaume, G., Doucet, P., Song, A. H., Lu, M. Y., Almagro-Pérez, C., Wagner, S. J.,… & Mahmood, F. (2024). Hest-1k: A dataset for spatial transcriptomics and histology image analysis. Advances in Neural Information Processing Systems, 37, 53798–53833.

[30] Zhang, A., Jaume, G., Vaidya, A., Ding, T., & Mahmood, F. (2025). Accelerating data processing and benchmarking of ai models for pathology. arXiv preprint arXiv:2502.06750.

[31] Bankhead, P., Loughrey, M. B., Fernández, J. A., Dombrowski, Y., McArt, D. G., Dunne, P. D., McQuaid, S., Gray, R. T., Murray, L. J., Coleman, H. G., James, J. A., Salto-Tellez, M., & Hamilton, P. W. (2017). QuPath: Open source software for digital pathology image analysis. Scientific reports, 7(1), 16878. 10.1038/s41598-017-17204-5

[32] Ding, T., Wagner, S. J., Song, A. H., Chen, R. J., Lu, M. Y., Zhang, A., Vaidya, A. J., Jaume, G., Shaban, M., Kim, A., Williamson, D. F. K., Robertson, H., Chen, B., Almagro-Pérez, C., Doucet, P., Sahai, S., Chen, C., Chen, C. S., Komura, D., Kawabe, A.,… Mahmood, F. (2025). A multimodal whole-slide foundation model for pathology. Nature medicine, 31(11), 3749–3761. 10.1038/s41591-025-03982-3

[33] Lurkin, A., Ducimetière, F., Vince, D. R., Decouvelaere, A. V., Cellier, D., Gilly, F. N., Salameire, D., Biron, P., de Laroche, G., Blay, J. Y., & Ray-Coquard, I. (2010). Epidemiological evaluation of concordance between initial diagnosis and central pathology review in a comprehensive and prospective series of sarcoma patients in the Rhone-Alpes region. BMC cancer, 10, 150. 10.1186/1471-2407-10-150

[34] Thway, K., & Fisher, C. (2009). Histopathological diagnostic discrepancies in soft tissue tumours referred to a specialist centre. Sarcoma, 2009, 741975. 10.1155/2009/741975

[35] Al-Ibraheemi, A., & Folpe, A. L. (2016). Voluntary Second Opinions in Pediatric Bone and Soft Tissue Pathology: A Retrospective Review of 1601 Cases From a Single Mesenchymal Tumor Consultation Service. International journal of surgical pathology, 24(8), 685–691. 10.1177/1066896916657591

[36] Perrier, L., Rascle, P., Morelle, M., Toulmonde, M., Ranchere Vince, D., Le Cesne, A., Terrier, P., Neuville, A., Meeus, P., Farsi, F., Ducimetière, F., Blay, J. Y., Ray Coquard, I., & Coindre, J. M. (2018). The cost-saving effect of centralized histological reviews with soft tissue and visceral sarcomas, GIST, and desmoid tumors: The experiences of the pathologists of the French Sarcoma Group. PloS one, 13(4), e0193330. 10.1371/journal.pone.0193330

[37] Kawai, A., Yoshida, A., Shimoi, T., Kobayashi, E., Yonemori, K., Ogura, K., Iwata, S., & Toshirou, N. (2024). Histological diagnostic discrepancy and its clinical impact in bone and soft tissue tumors referred to a sarcoma center. Cancer science, 115(8), 2831–2838. 10.1111/cas.16211

[38] Eckardt, M. A., Siena, N. M., Copeland, A. R., McCaw, T. R., Shehata, M. S., Lofftus, S. Y., Graham, D. S., Dao, H. B., Singh, A. S., Chmielowski, B., Federman, N., Kadera, B. E., Kalbasi, A., Bernthal, N. M., Eilber, F. R., Dry, S. M., Nelson, S. D., Eilber, F. C., & Crompton, J. G. (2025). Dedicated review of sarcoma pathology is necessary for corroborative diagnosis in nearly one half of referred patients. Surgery, 188, 109610. 10.1016/j.surg.2025.109610

[39] Lu, M. Y., Chen, T. Y., Williamson, D. F. K., Zhao, M., Shady, M., Lipkova, J., & Mahmood, F. (2021). AI-based pathology predicts origins for cancers of unknown primary. Nature, 594(7861), 106–110. 10.1038/s41586-021-03512-4

[40] Mantilla, J. G., Ricciotti, R. W., Chen, E. Y., Liu, Y. J., & Hoch, B. L. (2019). Amplification of DNA damage-inducible transcript 3 (DDIT3) is associated with myxoid liposarcoma-like morphology and homologous lipoblastic differentiation in dedifferentiated liposarcoma. Modern pathology: an official journal of the United States and Canadian Academy of Pathology, Inc, 32(4), 585–592. 10.1038/s41379-018-0171-y

[41] Ylagan, L. R., & Bhalla, S. (2001). Fine needle aspiration cytology of a dedifferentiated liposarcoma: report of a case with histologic and immunohistochemical follow-up. Acta cytologica, 45(4), 641–644. 10.1159/000327880

[42] Schaefer, I. M., & Gronchi, A. (2022). WHO Pathology: Highlights of the 2020 Sarcoma Update. Surgical oncology clinics of North America, 31(3), 321–340. 10.1016/j.soc.2022.03.001

[43] Gjorgova Gjeorgjievski, S., Thway, K., Dermawan, J. K., John, I., Fisher, C., Rubin, B. P., Jenkins, S., Thangaiah, J. J., Folpe, A. L., & Fritchie, K. J. (2022). Pleomorphic Liposarcoma: A Series of 120 Cases With Emphasis on Morphologic Variants. The American journal of surgical pathology, 46(12), 1700–1705. 10.1097/PAS.0000000000001962

[44] Qiu, G., Zhou, X., Shang, Z., Wang, Y., & Hai, R. (2025). Primary lipomatous tumor/well-differentiated liposarcoma of the breast with invasion of the pectoralis major muscle: A case report. International journal of surgery case reports, 133, 111680. 10.1016/j.ijscr.2025.111680

[45] Yamamoto, T., Minami, R., Ohbayashi, C., & Inaba, M. (2002). Epithelioid leiomyosarcoma of the external deep soft tissue. Archives of pathology & laboratory medicine, 126(4), 468–470. 10.5858/2002-126-0468-ELOTED

[46] Yang, Q., Madueke-Laveaux, O. S., Cun, H., Wlodarczyk, M., Garcia, N., Carvalho, K. C., & Al-Hendy, A. (2024). Comprehensive Review of Uterine Leiomyosarcoma: Pathogenesis, Diagnosis, Prognosis, and Targeted Therapy. Cells, 13(13), 1106. 10.3390/cells13131106

[47] Magro, G., Broggi, G., Angelico, G., Puzzo, L., Vecchio, G. M., Virzì, V., Salvatorelli, L., & Ruggieri, M. (2022). Practical Approach to Histological Diagnosis of Peripheral Nerve Sheath Tumors: An Update. Diagnostics (Basel, Switzerland), 12(6), 1463. 10.3390/diagnostics12061463

[48] Knight, S. W. E., Knight, T. E., Santiago, T., Murphy, A. J., & Abdelhafeez, A. H. (2022). Malignant Peripheral Nerve Sheath Tumors-A Comprehensive Review of Pathophysiology, Diagnosis, and Multidisciplinary Management. Children (Basel, Switzerland), 9(1), 38. 10.3390/children9010038

[49] Chang F. (2006). Desmoplastic small round cell tumors: cytologic, histologic, and immunohistochemical features. Archives of pathology & laboratory medicine, 130(5), 728–732. 10.5858/2006-130-728-DSRCTC

[50] Shakked, R. J., Geller, D. S., Gorlick, R., & Dorfman, H. D. (2012). Mesenchymal chondrosarcoma: clinicopathologic study of 20 cases. Archives of pathology & laboratory medicine, 136(1), 61–75. 10.5858/arpa.2010-0362-OA

[51] Li, C., Krasniqi, F., Donners, R., Kettelhack, C., & Krieg, A. H. (2024). Synovial sarcoma: the misdiagnosed sarcoma. EFORT open reviews, 9(3), 190–201. 10.1530/EOR-23-0193

[52] Hisaoka, M., Ishida, T., Kuo, T. T., Matsuyama, A., Imamura, T., Nishida, K., Kuroda, H., Inayama, Y., Oshiro, H., Kobayashi, H., Nakajima, T., Fukuda, T., Ae, K., & Hashimoto, H. (2008). Clear cell sarcoma of soft tissue: a clinicopathologic, immunohistochemical, and molecular analysis of 33 cases. The American journal of surgical pathology, 32(3), 452–460. 10.1097/PAS.0b013e31814b18fb

[53] Kosemehmetoglu, K., & Folpe, A. L. (2010). Clear cell sarcoma of tendons and aponeuroses, and osteoclast-rich tumour of the gastrointestinal tract with features resembling clear cell sarcoma of soft parts: a review and update. Journal of clinical pathology, 63(5), 416–423. 10.1136/jcp.2008.057471

[54] Li, Y., Cao, G., Tao, X., Guo, J., Wu, S., & Tao, Y. (2019). Clinicopathologic features of epithelioid sarcoma: report of seventeen cases and review of literature. International journal of clinical and experimental pathology, 12(8), 3042–3048.

[55] Guillou, L., Wadden, C., Coindre, J. M., Krausz, T., & Fletcher, C. D. (1997). “Proximal-type” epithelioid sarcoma, a distinctive aggressive neoplasm showing rhabdoid features. Clinicopathologic, immunohistochemical, and ultrastructural study of a series. The American journal of surgical pathology, 21(2), 130–146. 10.1097/00000478-199702000-00002

[56] Durfee, R. A., Mohammed, M., & Luu, H. H. (2016). Review of Osteosarcoma and Current Management. Rheumatology and therapy, 3(2), 221–243. 10.1007/s40744-016-0046-y

[57] Thway, K., & Fisher, C. (2021). Undifferentiated and dedifferentiated soft tissue neoplasms: Immunohistochemical surrogates for differential diagnosis. Seminars in diagnostic pathology, 38(6), 170–186. 10.1053/j.semdp.2021.09.005

[58] Janowczyk, A., Basavanhally, A., & Madabhushi, A. (2017). Stain Normalization using Sparse AutoEncoders (StaNoSA): Application to digital pathology. Computerized medical imaging and graphics: the official journal of the Computerized Medical Imaging Society, 57, 50–61. 10.1016/j.compmedimag.2016.05.003

[59] Xu, J., Xiang, L., Liu, Q., Gilmore, H., Wu, J., Tang, J., & Madabhushi, A. (2016). Stacked Sparse Autoencoder (SSAE) for Nuclei Detection on Breast Cancer Histopathology Images. IEEE transactions on medical imaging, 35(1), 119–130. 10.1109/TMI.2015.2458702

[60] Le NM, Shen C, Patel N, et al. Learning biologically relevant features in a pathology foundation model using sparse autoencoders. arXiv:2407.10785. 2024.

[61] Diao, J. A., Wang, J. K., Chui, W. F., Mountain, V., Gullapally, S. C., Srinivasan, R., Mitchell, R. N., Glass, B., Hoffman, S., Rao, S. K., Maheshwari, C., Lahiri, A., Prakash, A., McLoughlin, R., Kerner, J. K., Resnick, M. B., Montalto, M. C., Khosla, A., Wapinski, I. N., Beck, A. H.,… Taylor-Weiner, A. (2021). Human-interpretable image features derived from densely mapped cancer pathology slides predict diverse molecular phenotypes. Nature communications, 12(1), 1613. 10.1038/s41467-021-21896-9

[62] Xu, S., Hou, J., & Chen, H. (2025, September). Explain Any Pathological Concept: Discovering Hierarchical Explanations for Pathology Foundation Models. In International Conference on Medical Image Computing and Computer-Assisted Intervention (pp. 216–225). Cham: Springer Nature Switzerland.

[63] Geirhos, R., Jacobsen, J. H., Michaelis, C., Zemel, R., Brendel, W., Bethge, M., & Wichmann, F. A. (2020). Shortcut learning in deep neural networks. Nature Machine Intelligence, 2(11), 665–673.

[64] Ong Ly, C., Unnikrishnan, B., Tadic, T., Patel, T., Duhamel, J., Kandel, S., Moayedi, Y., Brudno, M., Hope, A., Ross, H., & McIntosh, C. (2024). Shortcut learning in medical AI hinders generalization: method for estimating AI model generalization without external data. NPJ digital medicine, 7(1), 124. 10.1038/s41746-024-01118-4

[65] Homeyer, A., Geißler, C., Schwen, L. O., Zakrzewski, F., Evans, T., Strohmenger, K., Westphal, M., Bülow, R. D., Kargl, M., Karjauv, A., Munné-Bertran, I., Retzlaff, C. O., Romero-López, A., Sołtysiński, T., Plass, M., Carvalho, R., Steinbach, P., Lan, Y. C., Bouteldja, N., Haber, D.,… Zerbe, N. (2022). Recommendations on compiling test datasets for evaluating artificial intelligence solutions in pathology. Modern pathology: an official journal of the United States and Canadian Academy of Pathology, Inc, 35(12), 1759–1769. 10.1038/s41379-022-01147-y

